# Efficacy of a fixed-ratio combination of glucagon-like peptide-1 receptor agonist and basal insulin therapy for the treatment of type 2 diabetes: a protocol for systematic review and meta-analysis of randomized clinical trials

**DOI:** 10.1101/2024.07.18.24310630

**Authors:** Mansur A. Ramalan, Musa Baba Maiyaki, Ibrahim D Gezawa, Andrew E Uloko

**Affiliations:** Department of Internal Medicine, Aminu Kano Teaching Hospital, Kano, Kano State, Nigeria; Department of Internal Medicine, Bayero University, Kano, Kano State, Nigeria; Africa Center of Excellence for Population Health and Policy Bayero University Kano, Kano State, Nigeria

**Author notes:** Corresponding author (MAR). **Author contribution Mansur Aliyu Ramalan**, Conceptualization, Methodology, Project administration, Writing – original draft, Writing – review & editing, **Ibrahim D Gezawa**, Writing – original draft, Writing – review & editing, **Andrew Uloko** Writing – original draft, Writing – review & editing, **Musa Babamaiyaki**, Methodology, Writing – review & editing. Prospero registration number CRD42022363619.

## Abstract

**Background:** Blood glucose control is a major goal of management demonstrated by many guidelines and results of clinical trials. Achieving good glycaemia is associated with reduction of the risks of cardiovascular disease. Glucagon like peptide 1 receptor agonists (GLP-1 RAs) are now important parts of many guidelines for the management of diabetes mellitus because of their cardiovascular benefit. Basal insulin in have now been combined with GLP-1 RAs in fixed ration combinations in single pens. We aim to conduct a meta-analysis and systematic review of randomized control clinical trials the evaluate the therapeutic efficacy of glucagon-like peptide-1 receptor agonists combined with basal insulin therapy in treating type 2 diabetes, using a fixed-ratio combination of the two.

**Method and data analysis:** It is anticipated that a systematic search of the literature will be conducted on the following electronic databases will be searched: The Cochrane Library, PubMed, Google Scholar, and EMBASE. We plan to include only randomized control clinical trials on GLP 1RA, fixed ratio combination of basal insulin and GLP 1RA and basal insulins alone. A validated study design-specific critical appraisal tool will be used to assess the risk of bias in individual studies. The systematic review will be performed in accordance with the JBI guideline for prevalence and incidence review. The review will be reporting following the Preferred Reporting Items for Systematic Review and Meta-Analyses (PRISMA) guideline.

## Introduction

In patients with T2DM, blood glucose control is a major goal of management demonstrated by many guidelines and results of clinical trials.(1–3) It has been reported in the literature that micro and macro vascular risks increased with poor glycaemic control. The UKPDS for instance found that improved control of blood glucose or blood pressure reduced the risk of major diabetic eye disease by one quarter, serious deterioration of vision by nearly one half, early kidney damage by one third, strokes by one third, and death from diabetes-related causes by one third.(3)

Over the past few decades, the cardiovascular benefits and lower risks of hypoglycaemia of the Glucagon like peptide 1 receptor agonists (GLP-1 RAs) has made them a major part of many guidelines for the management of diabetes mellitus.(4,5) Basal insulins on the hand have been shown to be highly effective in glycaemic control of patients with T2DM.(6) They reduce fasting blood glucose levels and improve glycaemia but are associated with higher risks of hypoglycaemia compared to the GLP-1 RAs.

Recently the potential benefit of using the combination of GLP-1 RAs and basal insulin in the pharmacological treatment of type 2 diabetes mellitus has been shown to improve glycemic control. (7,8)

There are several combinations of GLP-1 RAs and basal insulins in fixed ratio combinations available for use in a single injection pen currently available. (9)

The potential benefits of combining GLP-1RAs with basal insulin in fixed ratio combinations is to Improve glycaemia, while reducing the risks of hypoglycaemia and lower the risks of weight gain.(9)

This protocol intends to review the therapeutic efficacy, safety and cardiovascular benefit of the fixed ratio combinations of the GLP-1 RAs and basal insulins in comparison with the GLP-1 RAs alone or basal insulins alone.

## Aim and objectives

This study aims to conduct a systematic review and meta-analysis of randomized controlled clinical trials with the aim of evaluating the therapeutic efficacy of glucagon-like peptide-1 receptor agonists combined with basal insulin therapy in treating type 2 diabetes, using a fixed-ratio combination of the two.

The secondary objectives of the systematic review will include:

1. to determine the cardiovascular outcome of a fixed-ratio combination of glucagon-like peptide-1 receptor agonist versus basal insulin alone or in combination with prandial insulin therapy for the treatment of type 2 diabetes
2. to determine the incidence of adverse events in patients with type 2 diabetes on a fixed-ratio combination of glucagon-like peptide-1 receptor agonist versus basal insulin alone or in combination with prandial insulin
3. to compare the effects of a fixed-ratio combination of glucagon-like peptide-1 receptor agonist versus basal insulin alone or in combination with prandial insulin on weight and hypoglycemia in patients with type 2 diabetes.

## Method

### Registration and reporting of the review findings

The protocol for this review has been registered with Prospero with the registration number CRD42022363619. We will use PRISMA (Preferred Reporting Items for Systematic Reviews and Meta-Analyses)(10) to report the findings of this study. In order to conduct this systematic review, we will follow the guidelines set by the Joanna Briggs Institute (JBI) for systematic reviews.(11)

### Study eligibility

Population, intervention, comparison, and outcome (PICO) protocols will be used to evaluate study eligibility.

#### Population

Randomized controlled trials conducted on adult T2DM patients.

#### Intervention

GLP-1RA and basal insulin, or GLP-1RA alone or basal insulin alone.

#### Comparison

T2DM patients using GLP-1RA and basal insulin will be compared to GLP-1RA alone or basal insulin alone (comparator).

#### Outcome

Reduction in HbA1c from baseline, reduction in fasting plasma glucose levels, hypoglycemia, and weight gain.

### Search strategy

The Preferred Reporting Items for Systematic Reviews and Metanalyses checklist will be utilized for the systematic review of the literature. Searches of the literature will be conducted in the Cochrane Library, PubMed, Google Scholar, and EMBASE. The research will be randomized control trials only. There will be no language restrictions on the studies and articles selected for the review. The search strategy will include terms for GLP 1 agonists and basal insulin fixed-dose combination, GLP 1 agonists, basal insulin, basal-bolus insulin, prandial insulin, cardiovascular benefits, adverse events, hypoglycemia, weight reduction, efficacy, and adherence.

### Population characteristics

1. Patients with type 2 diabetes who were initiated on the fixed ratio combination or switched to the fixed ratio combination for at least 12 weeks
2. Patients with type 2 diabetes either initiated on basal insulin or prandial insulin with or without basal insulin or switched to basal insulin or prandial insulin with or without basal insulin for at least 12 weeks
3. Outcome measures: therapeutic efficacy, cardiovascular outcome, adverse events and effects on weight. Basal insulin or prandial insulin with or without basal insulin or switched to basal insulin or prandial insulin with or without basal insulin for at least 12 weeks
4. Secondary outcome measures: Therapeutic efficacy, cardiovascular outcome, adverse events and effects on weight

### Research question

What is the therapeutic efficacy of GLP1 agonist and basal insulin fixed-dose combinations in the treatment of type 2 diabetes compared to GLP1 agonists or basal insulin alone?

What is the cardiovascular benefit of using GLP1 agonist and basal insulin fixed-dose combinations in the treatment of type 2 diabetes compared to GLP1 agonists or basal insulin alone?

What is the burden of adverse effects of GLP1 agonist and basal insulin fixed-dose combinations in the treatment of type 2 diabetes compared to GLP1 agonists or basal insulin alone?

### Eligibility criteria

#### Inclusion criteria

Randomized controlled clinical trials that provide estimates of efficacy, adverse events or cardiovascular outcomes and effects on weight.

##### Outcome index

therapeutic efficacy, cardiovascular outcome, adverse events and effects on weight.

#### Exclusion criteria

Qualitative studies, case reports, case series, case-control studies, conference proceedings, prospective and retrospective observational studies, descriptive and analytical cross-sectional studies. Expert opinions, and randomized control trials not reporting estimates of therapeutic efficacy, unpublished data, repeated publication of literature, or where the full text is not available. Or where the data cannot be extracted.

### Language

Only studies written in English or with an available English version, or with the possibility of translating the full article via Google Translate, will be included.

### Study period

We will focus on the past three decades; therefore, we will include studies conducted from 1^st^ January 1990 to 30^th^ of June 2023.

### Electronic searches

It is anticipated that the following electronic databases will be searched: The Cochrane Library, PubMed, Google Scholar, and EMBASE. The research will be randomized control trials only. There will be no language restrictions on studies and articles selected for the review. The search strategy will include terms for GLP 1 agonists and basal insulin fixed-dose combination, GLP 1 agonists, basal insulin, basal-bolus insulin, prandial insulin, cardiovascular benefits, adverse events, hypoglycemia, weight reduction, efficacy, and adherence. (See S1 Appendix). Terms representing similar concepts will be separated using the Boolean operator “OR” while terms representing different concepts will be separated using the Boolean operator “AND”. Where the full text of relevant studies cannot be retrieved, efforts will be made to request the full text from study corresponding authors.

The literature will be independently screened by 2 researchers to identify articles that meet the inclusion criteria. Firstly, according to the weight of the topic, review the topic summary to exclude irrelevant literature. Then read the full text to select literature according to the inclusion and exclusion criteria. Finally, two people will check the results, and if their opinions don’t agree, a third party will take the decision. The following contents will be extracted from the included literature: first author, year of publication, age of study subjects, study site, estimates of efficacy, adverse events, effects of weight and hypoglycaemia. The reasons for the exclusion of full-text articles will be given. PRISMA flow diagrams will be used to describe and summarize the search results and selection process. A table will be used to report the details of all the included studies.

### Other searches

A hand search will be conducted for unpublished studies/grey literature using the keywords found in the study through Google Scholar and gray literature databases such as WHO Library, OpenSIGLE, and Open Gray.

### Risk of bias

The risk of bias in the included studies will be assessed using the Joanna Briggs Institute (JBI) tool. We will describe any assessment of the risk of bias that may affect the cumulative evidence (e.g., publication bias, selective reporting within studies).

### Data extraction

The primary changes in HbA1c from baseline to the end of treatment. Changes in the levels of fasting plasma glucose from baseline to the end of the study, changes in the body weights of the participants and the proportion of participants reaching the HbA1c target <7.0% at the end of the study. Reported and documented hypoglycaemic episodes during the period of treatment.

### Strategy for data synthesis

A quantitative meta-analysis will be performed on the estimates of efficacy, adverse events, effects of weight and hypoglycaemia in patients with type 2 diabetes using Stata17.0. Pooled estimates of weighted means differences (WMDS) and 95% CI for continuous outcomes; changes in HbA1c, FPG, as well as the pooled risk ratios (RR) of dichotomous outcomes including the proportion of participants achieving HbA1c target values and the risks of hypoglycaemia will be calculated.

Heterogeneity will be assessed using the I^2^ statistic for each analysis, with the significance level set at P<0.05. If there is found to be significant heterogeneity between the studies, the random effects model will be used, or these studies will be divided into specific subgroups, based on different factors. The potential risks of publication bias will be evaluated by constructing funnel plots of primary outcomes with asymmetry assessed by Egger’s test.

A forest plot will be used to illustrate the distribution of the outcome and effect sizes obtained from each published study. In addition, publication bias will also be assessed, and a sensitivity analysis carried out.

### Analysis of subgroups or subsets

A sensitivity analysis comparing the intervention arm GLP-1 agonist and basal insulin with basal insulin therapy or basal-bolus insulin is planned. Subgroup analysis will also be conducted. The efficacy of individual FRC drugs will be reported using a forest plot.

### Assessment of certainty of evidence

To assess the quality of the synthesised findings, we will use the GRADE instrument and Gradepro Software. Studies included in the analysis will be assessed for their design, heterogeneity, consistency, directness, precision, and publication bias. A quality rating will be assigned to the evidence: high, moderate, low or very low.

## Discussion

The proposed study will provide the first insight into the comparative efficacy, safety, and adverse effects profile of the fixed ration combination of GLP-1RA and basal insulin in comparison to basal insulin alone or GLP1-RA alone. Also, the specific cardiovascular benefit of the fixed ration combination of the GLP-1 receptor agonist with basal insulin has not been completely elucidated although the cardiovascular benefit of GLP-1 RA has been well established.(9) The fixed-ratio combination of GLP1 RA and basal insulin is relatively new but in previous studies that compared with the basal-bolus insulin regimen, the combination of GLP-1RA and basal insulin displayed a similar benefit in lowering HbA1c with a 33% lower risk of hypoglycemia and 5.66 kg less body weight gain.(12)

The combination of basal insulin GLP -1 RA is a synergistic effect that is produced by the activity of the basal insulin principally suppressing fasting plasma glucose, and the GLP 1 RA that acts by suppressing the postprandial glucose excursions.

We expect that the study will be conducted using a systematic approach utilizing a well-defined protocol and following a standard algorithm for data collection and analysis. Among the potential limitations, heterogeneity in individual studies may be encountered due to the quality of the study designs, definitions used, or evaluation methods. A detailed narrative report will, however, be provided highlighting these discrepancies and providing suggestions for improving future research.

## Data Availability

The data is not yet available but will be provided when the process is completed

## Acknowledgments

Nil

Source of funding - Self funded

Supporting information

Appendix 1

PRISMA Check list

